# A modeling framework for translating discrete choice experiment results into cost-effectiveness estimates: an application to designing tailored and scalable HIV and contraceptive services for adolescents in South Africa

**DOI:** 10.1101/2022.09.08.22279581

**Authors:** Caroline Govathson, Lawrence C Long, Colin A Russell, Aneesa Moolla, Sophie Pascoe, Brooke E. Nichols

## Abstract

**Background:** Youth in South Africa are disproportionately affected by STIs, HIV, and unintended pregnancies. Despite this, their uptake of HIV and contraceptive services remains a challenge. South Africa urgently needs tailored, scalable interventions to address both HIV infection and early pregnancy prevention for young people. These interventions generally take years to design, implement, and evaluate, leaving a gap. To that end, we have developed a framework to translate the expected impact of facility-level attributes in increasing HIV/contraceptive service uptake for youth from a discrete choice experiment (DCE) into a cost effectiveness analysis (CEA).

**Methods:** We used a DCE (n=805) conducted in Gauteng, South Africa, which found that staff attitude, confidentiality, Wi-Fi, subsidized food, afternoon hours and youth-only services were preferred attributes of health services. Based on this we simulated uptake of services adapted for these preferences. We divided preferences into modifiable attributes that could readily be adapted (e.g. Wi-Fi), and non-modifiable (more nuanced attributes that are more challenging to cost and evaluate): staff attitude and estimated the incremental change in uptake of services using adapted services. Costs for modifiable preferences were estimated using data from two clinics in South Africa (2019 US$). We determined the incremental cost effectiveness ratio (ICER) of 15 intervention combinations, and report the results of interventions on the cost-effectiveness frontier.

**Results:** Greatest projected impact on uptake was from friendly and confidential services, both of which were considered non-modifiable (18.5% 95%CI:13.0-24.0%; 8.4% 95%CI:3.0-14.0% respectively). Modifiable factors on their own resulted in only small increases in expected uptake. (Food: 2.3% 95%CI:4.0%-9.00%; Wi-Fi: 3.0% 95%CI: -4.0%-10.0%; Youth only services: 0.3% 95%CI: -6.0%-7.0%; Afternoon services: 0.8% 95%CI: -6.0%-7.0%). The order of interventions on the cost-effectiveness frontier are Wi-Fi and youth-only services (ICER US$7.01-US$9.78), Wi-Fi, youth-only services and food (ICER US$9.32 - US$10.45), followed by Wi-Fi, youth-only services and extended afternoon hours (ICER US$14.46 – US$43.63)

**Conclusion:** Combining DCE results and costing analyses within a modelling framework provides an innovative way to inform decisions on effective resource utilisation. Modifiable preferences, such as Wi-Fi provision, youth only services and subsidized food, have potential to cost-effectively increase the proportion of youth accessing HIV and contraceptive services.

## Introduction

Existing models of care have failed to adequately address the gap in sexual health needs, access to- and uptake of-HIV and contraceptive services among adolescents in South Africa (1). The rate of unintended pregnancy among adolescents remains high, with about a quarter of adolescent girls and young women (AWYG) giving birth before age 20 (2)(3). According to UNAIDS, the incidence rate for HIV infection among adolescents aged 10-19 was 54 per 1000 population in 2018 in South Africa with adolescent girls disproportionally affected(4). Despite the high prevalence of HIV and pregnancy among adolescents, uptake of HIV and contraceptive services in this age group remains a challenge. The 2016 South African demographic health survey revealed that 31% of girls aged 15-19 years and 28% of AGYW aged 20-24 years had unmet contraceptive needs(5). This underscores a critical need to increase adolescent access to both HIV and contraceptive services.

Many solutions have been offered to increase uptake of HIV and contraceptive services by adolescents including “youth friendly clinics” and school health programs with limited success (1,6,7). Children 12 years and older have the right to reproductive health, including access to contraceptives and HIV/AIDS testing without consent of a parent or guardian (8). There however remain many barriers to adolescents’ access and uptake of reproductive health services (9–11). South Africa urgently needs tailored, scalable interventions to address both HIV infection and early pregnancy prevention for young people, in particular for young women. Decisions on how to tailor and efficiently scale interventions with regards to cost and uptake are often difficult to make due to limited data on youth preferences, impact and cost of different interventions.

Discrete choice experiments (DCE’s) are a study design increasingly used in health economics to explore the relative importance of different attributes that may influence decision to access and utilize certain goods, services or programs (12,13). They can be used to understand whether particular attributes can predict uptake of services and the relative importance of certain attributes to this uptake (14). The results from a DCE can help policy makers understand the impact of making certain changes to current services on service acceptability or uptake. Recent research has been conducted to determine the preferences of school going youth for HIV and contraceptive services through the use of a DCE(15). Key service characteristics including staff attitude, confidentiality and value-added services like availability of Wi-Fi, food and youth-only waiting areas were found to be key attributes affecting the decision to access care. The cost and expected effectiveness of altering these key attributes to improve the likelihood that the youth access health services is, however, unknown.

When designing interventions using preference data from a DCE, information on incremental uptake and how much it costs to implement these interventions is essential(16,17). This information allows us to compare the different interventions and select only the most cost-effective, scalable interventions for trialing. Traditionally trial data, stakeholder opinions, observational data of comparable scenarios are used with some underlying assumptions to predict uptake and cost(16–19) In our work we describe an approach that can be used to close the gap between the DCE methodology and implementation or trialing of different targeted programs.

In order to guide future trials or implementation, we used the results of the DCE with school going youth (15) to model the expected costs and impact (in terms of increase in the proportion of youth accessing health services) of attributes of health services. We used coefficients of expected uptake of HIV and contraceptive services from a recently conducted DCE (20), as an estimated outcome measure as part of a cost-effectiveness analysis. This information can be used to determine which interventions or combination of interventions should take priority within limited budgets.

## Methods

This work builds on a discrete choice experiment (DCE) that was conducted to estimate preferences of school going adolescents for accessing HIV and contraceptive services(15). The DCE was conducted in 10 high schools situated in neighbourhoods of varying socio-economic status (SES) in Gauteng South Africa between 07/2018-09/2019 (**Figure 1**). A total of 805 students completed the survey for the adolescent DCE study. Over two thirds (68%) of the participants were female, and two-thirds (66%) of the students who participated were aged 15-17 years. Details of the DCE will be published elsewhere.

**Figure 1:**
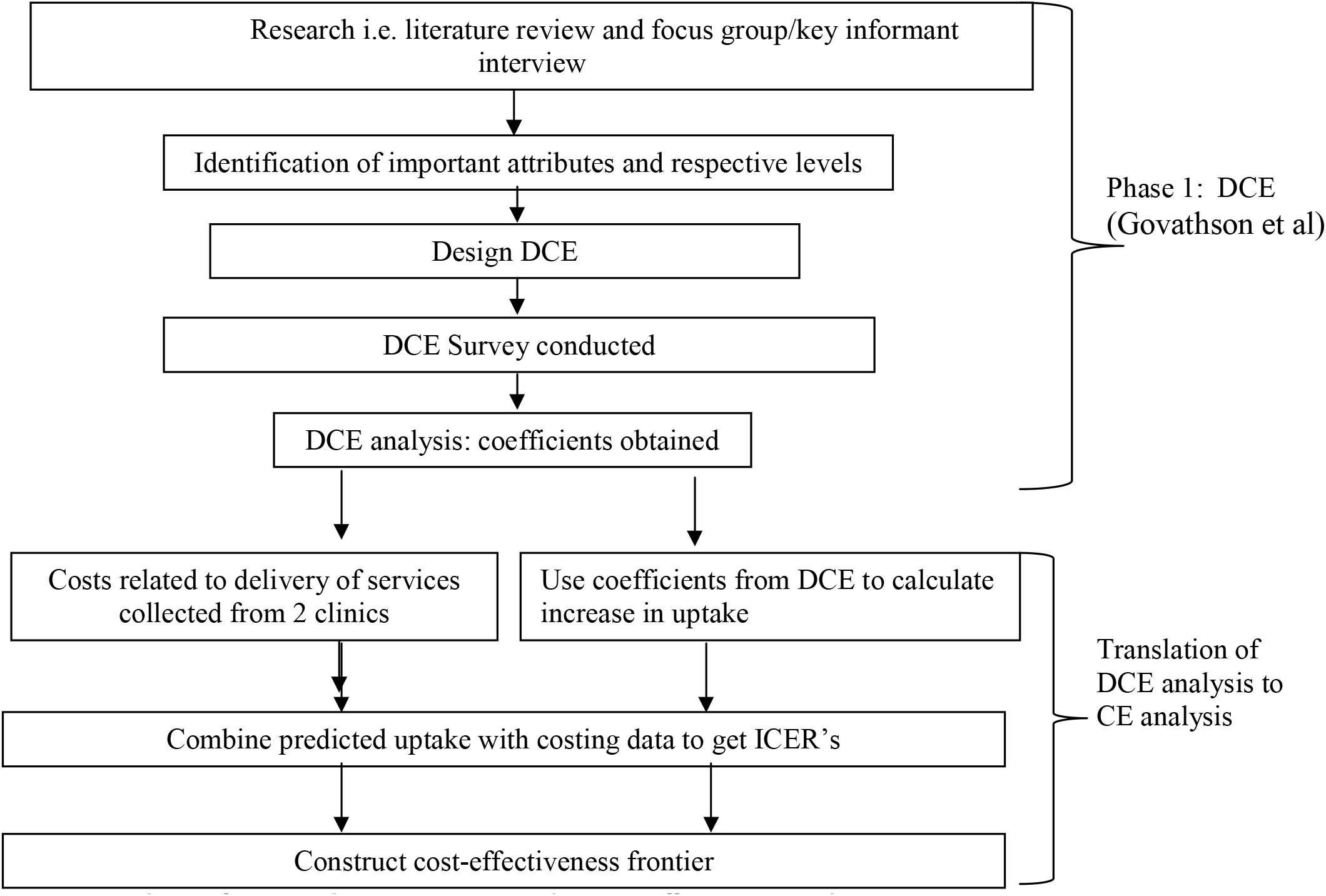
Translation of Discrete Choice Experiment results to cost-effectiveness results

From the above study, six factors were found to be facilitators for uptake of HIV and contraceptive services (15). These were the six attributes and levels that showed significant change in probability of a choice being selected. Staff attitude and confidentiality were key issues affecting students’ decisions to access HIV and contraceptive services. Access to subsidized food, youth-only waiting areas, afternoon services and presence of free Wi-Fi were the other preferences that were significantly associated with choice in the DCE. We used these results to identify combinations of interventions that could be used to increase uptake of HIV and contraceptive services by youths. The attributes were categorised into modifiable (easier to implement, and to cost and evaluate): subsidized food, Wi-Fi, youth-only services and afternoon services (after school hours); and non-modifiable (more nuanced attributes that are more challenging to cost and evaluate): friendliness and confidentiality. We looked at the potential increase in service uptake across the full set of mutually exclusive combinations of all six of these factors (Table 1). The complete cost-effectiveness analysis was, however, only done for the four factors that could be reasonably costed in a generalizable way. These four factors were placed into all possible combinations of between one and four attributes. This resulted in 15 different modifiable mutually exclusive intervention combinations that could be evaluated to see if they are cost effective options.

**Table 1.**
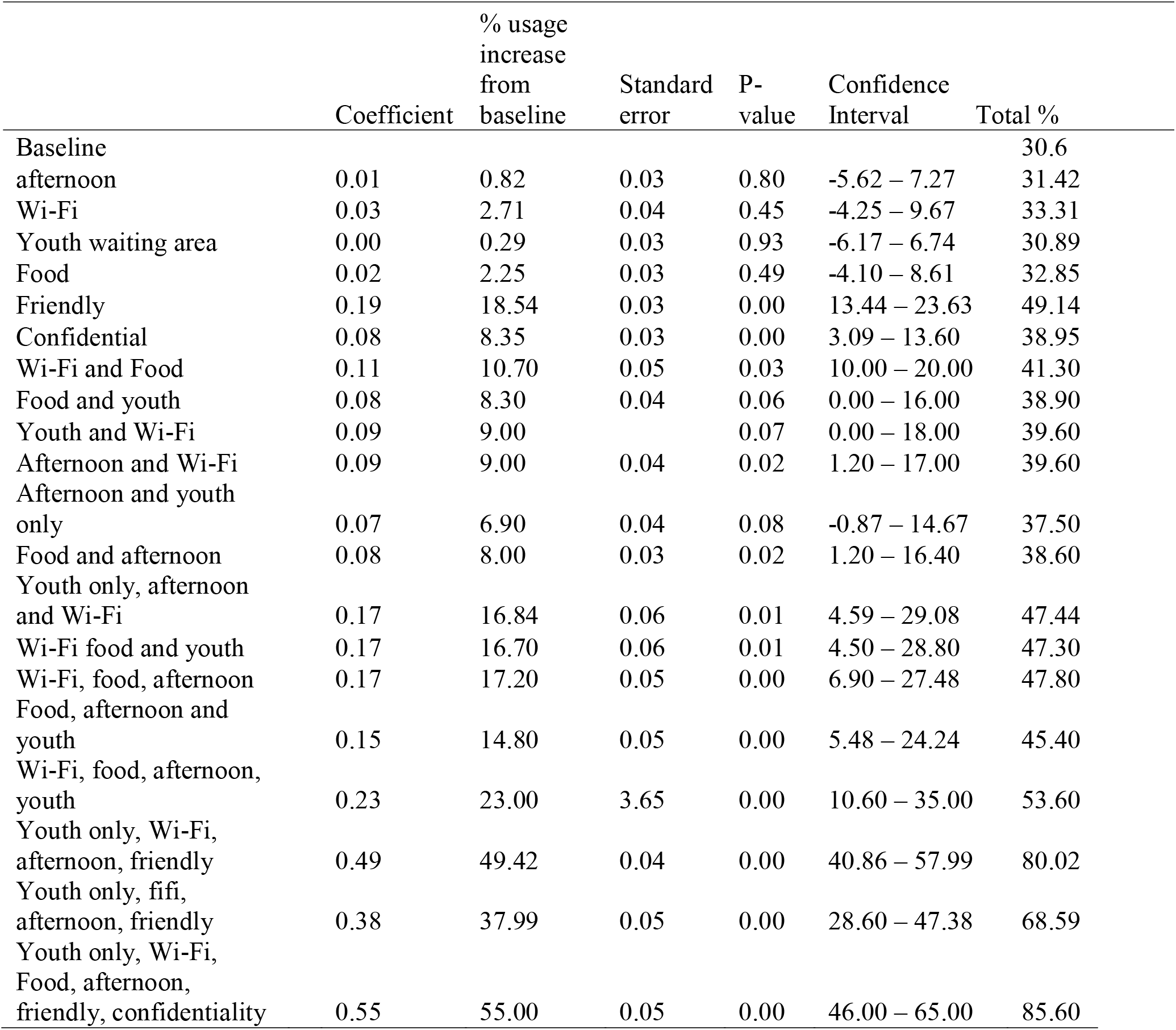
Uptake of HIV and contraceptive services projections for intervention scenarios

### Translation of DCE results to uptake

We used coefficients from the DCE (15) to estimate the expected increase in uptake of HIV and contraceptive services as the outcome measure for a cost-effectiveness analysis. We defined the baseline to be the set of attributes that most closely reflects current practice (Location: clinic, time: morning, staff attitude: unfriendly, confidentiality: none, incentives: none and provision of all health services). To estimate uptake of services, we calculated change of probability of uptake of using services from baseline levels when changes are made to the next level of each attribute (i.e. the incremental change in uptake). We modelled attributes, which had shown a statistically significant effect on the probability of an attribute being chosen in the DCE.

The logit probability of choosing alternative i rather than alternative j is given by the equation:

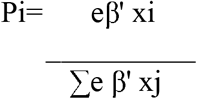

Using this equation (21), the change in the probability of choosing to use HIV and contraceptive services given the baseline levels say, clinic, and morning services is then (provided all other attributes remain equal) given by the above equation.

### Costs

Costs related to the delivery of a modified package of clinic interventions delivered with the aim of increasing the utilization of HIV and contraceptive services were estimated from a health system perspective.

Baseline costs were estimated from cost data collected from two clinics (one government primary health clinic and one South African non-profit organization providing Comprehensive PHC services with DOH support) through an external study focused on the costing of primary health services (22,23). The methods for the costing and the description of these primary health clinics are elsewhere (22–24). For each of the intervention scenarios, only the additional costs associated with the modification were calculated. The additional costs associated with the modification were shared among the additional adolescent patients expected to utilize the clinic following the changes.

The changes in cost in categories of assets, overheads, staff (clinical and non-clinical) and supplies for each package of interventions were calculated based on a number of assumptions regarding how these service modifications would be implemented and current local costs of implementing those changes. We used available cost analysis data and published data on interventions that have been implemented(23,25,26). All costs for the interventions were allocated to the additional adolescent patients and therefore division of total additional costs was by the number of extra adolescent patients.

Utilities such as electricity, water and sewage costs were allocated based on the total number of monthly visits. Staff costs for the afternoon and for youth only services were calculated based on number of patients seen and period of time the facility would be open in the afternoon. Food costs were calculated for all adolescents but the total cost was billed only to the additional adolescent patients so that the incremental cost included the cost of food to all adolescents. Extra space for youth-only services or afternoon services was calculated using average size counselling rooms.

We calculated a cost per additional adolescent patient expected to utilize HIV and contraceptive services as a consequence of the clinic modifications, as well as the total costs required at the facility level to implement the intervention. All costs were updated to 2019 South African Rand (ZAR) and converted to USD using the mid-year exchange rate 1 USD = 14 ZAR (27). Analysis of costs was done using Microsoft Excel.

### Cost-effectiveness ratios

In order to generate incremental cost-effectiveness ratios, the predicted probability analysis was combined with costing data for selected intervention scenarios. All intervention scenario costs and their respective expected effectiveness were incremental from a baseline cost and effectiveness of zero (e.g. no clinic modifications). We compared the new strategies (Intervention B) and the cost and outcome of these against the baseline standard of care (Intervention A) (i.e. Cost of Intervention B – Cost of intervention A)/ (Effectiveness of intervention B – Effectiveness of intervention A))(28). Those strategies that cost more and did not increase uptake of services were considered ‘dominated’ (e.g. costlier and less or equally effective) and therefore removed from the comparison. When the intervention increased the uptake of the services and was also less costly, we called it the “dominant” intervention. For interventions that increased uptake and also the cost, an ICER was calculated to compare the interventions, or a cost per additional youth expected to access services. To determine the most cost-effective strategies we used the efficiency frontier approach. A cost-effectiveness frontier representing the efficient combination of interventions of the 15 strategies assessed was constructed. Interventions below the frontier are not considered cost-effective at any threshold(29).

In the incremental analysis, strategies were compared until ultimately, only the most cost-effective strategies were left and these were ordered according to their cost-effectiveness ratio, lowest to highest, referencing which strategies should be adopted in incremental order dependent on the availability of resources(28,29). In order to determine how sensitive our results were to the specific costs at an individual clinic, we repeated the above analysis for two different clinics and their respective costs.

### Ethics

As this was a modelling study based on results from published studies that did not involve human subjects, approval from an institutional review board was not necessary.

## Results

### Preferences (Results from the DCE)

Results from the DCE that informed this work showed that the most preferred service characteristics that facilitated HIV and contraceptive service uptake by adolescents were friendly health care providers and private and confidential services. Students preferred accessing services in the afternoon (after school hours) compared to the morning. Youth only services, Wi-Fi and access to food were likely to increase the probability of students utilising services. Students preferred the more comprehensive package of services (family planning and contraceptive services) or all health services as opposed to services that only provided condoms or HIV counselling and Testing (HCT) (20)

### Uptake projections

The results from the simulations conducted here indicate that uptake of HIV and contraceptive services may increase as different service attributes are added to the baseline standard of care. Non-modifiable/difficult to modify factors such as ensuring friendly healthcare providers and confidentiality of services were projected to have the largest impact on uptake (an increase of 18.5%; (95%CI 13.0%-24.0%); and 8.4%; (95%CI 3.0%-14.0% respectively)). Combining these non-modifiable preferences with three modifiable preferences (Wi-Fi, food, youth only services and afternoon services) projected a statistically significant increase in uptake by 55.0% (95%CI 46.0%-65.0%) from baseline (**Figure 2**).

**Figure 2:**
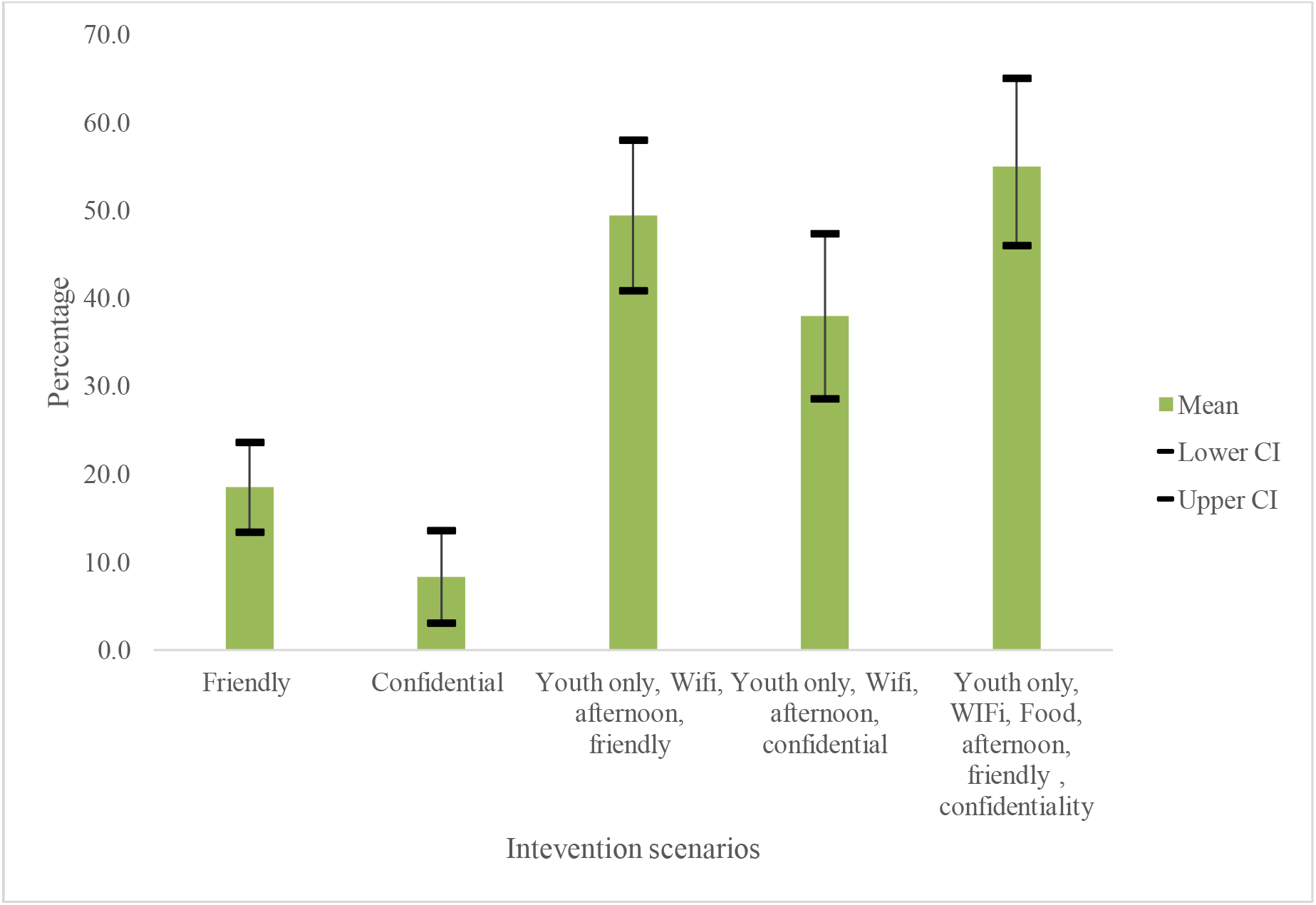
Uptake of HIV and contraceptive services projections for scenarios that include at least one non-modifiable attribute

For the final cost-effectiveness analysis, only modifiable attributes were included. As individual interventions, provision of afternoon services, or youth friendly services or Wi-Fi only or food only did not yield a statistically significant increase in rates of uptake of services. The least effective strategy in terms of additional youth expected to use HIV and contraceptive services were, in ascending order of efficacy, youth only services, afternoon services, access to subsidized food and provision of Wi-Fi separately. The combination of all four attributes, however, (Wi-Fi, food, afternoon services and youth) provided the highest increase in uptake of 23.0% (95%CI 11.0%-35.0%) (**Figure 3**). This was followed by different combinations of just three attributes which showed that uptake might be increased by roughly 17.0% for any combination of the three attributes (**Table 1**). Combining only two preferred modifiable attributes might also increase uptake by between 7.0% and 11.0% depending on combination of preferred attributes (**Table 1**). The number of adolescents utilising HIV and contraceptive services at baseline were higher at clinic B than at clinic A, therefore as a fraction, clinic A had more additional adolescents utilising services.

**Figure 3:**
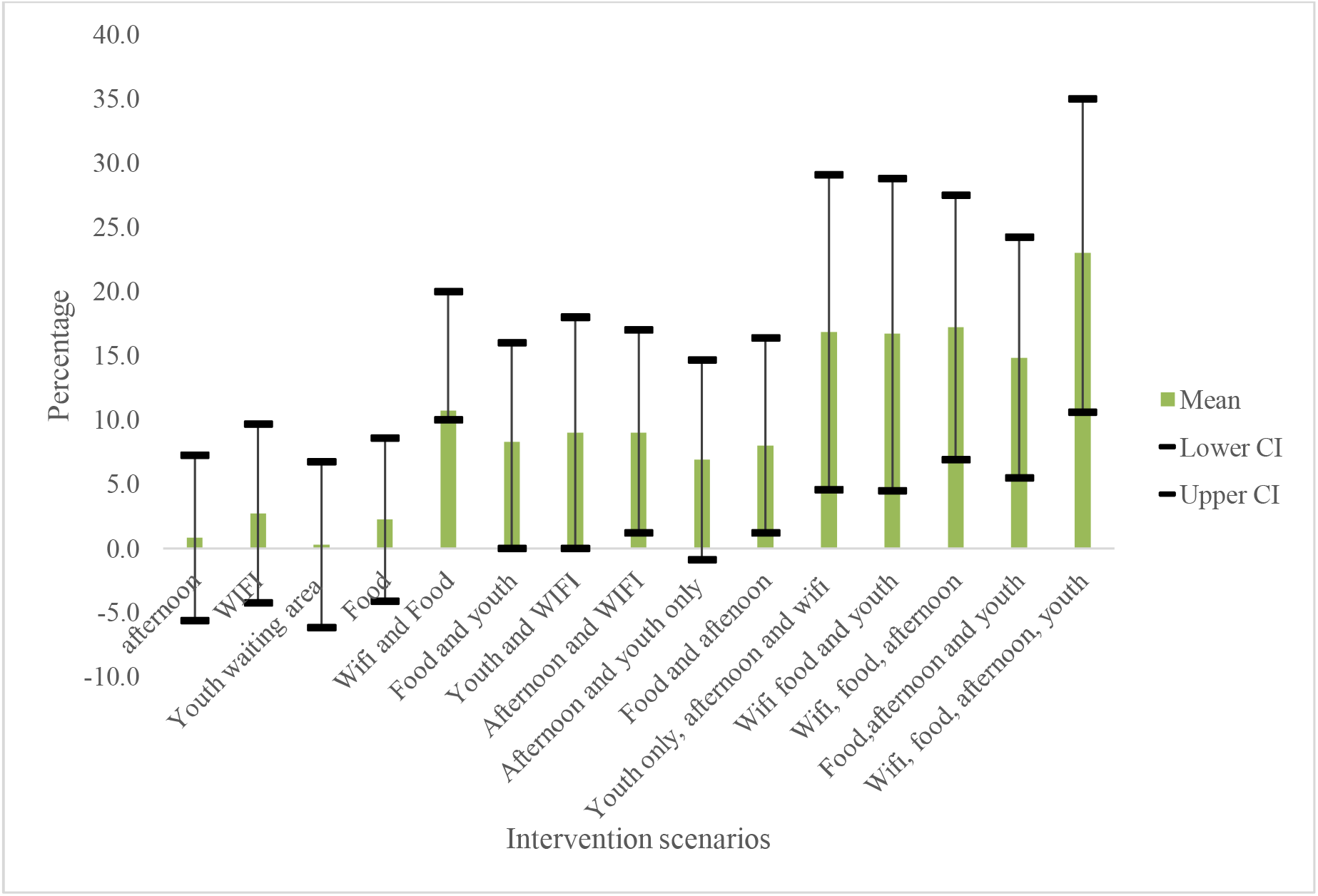
Uptake of HIV and contraceptive services projections for modifiable intervention scenarios

### Cost and cost-effectiveness

The cost per additional adolescent utilising services varied between the clinics depending on which interventions or package of interventions might be implemented. Clinic A costs ranged from $10.20 to 202.90 (**Table 2**) and Clinic B from $7.00 to $116.50 for clinic B (**Table 3**).

**Table 2:** Clinic A: Cost-effectiveness of interventions to increase youth uptake of HIV and contraceptive services

| Number | Additional youths | New total youths | % | Cost per patient | Total Cost | ICER |
| --- | --- | --- | --- | --- | --- | --- |
| Wi-Fi | 41 | 1572 | 2.7 | \$12.6 | \$516.7 | Dominated |
| youth only | 4 | 1535 | 0.3 | \$202.9 | \$811.7 | Dominated |
| afternoon | 13 | 1544 | 0.8 | \$78.5 | \$1,020.2 | Dominated |
| Food | 34 | 1565 | 2.3 | \$31.5 | \$1,072.3 | Dominated |
| Wi-Fi, youth only | 138 | 1669 | 9.0 | \$10.6 | \$1,465.6 | \$9.8 |
| Wi-Fi, food | 168 | 1699 | 10.7 | \$10.2 | \$1,706.0 | Weakly dominated |
| food, youth only | 122 | 1653 | 8.3 | \$15.2 | \$1,858.6 | Dominated |
| Wi-Fi, afternoon | 142 | 1673 | 9.3 | \$18.3 | \$2,603.3 | Weakly dominated |
| Wi-Fi, food, youth only | 260 | 1791 | 17 | \$10.5 | \$2,740.0 | \$10.5 |
| afternoon, youth only | 106 | 1637 | 6.9 | \$27.0 | \$2,864.9 | Dominated |
| Wi-Fi, afternoon, youth only | 230 | 1761 | 17.0 | \$13.7 | \$3,147.3 | Dominated |
| food, afternoon | 138 | 1669 | 9.0 | \$24.9 | \$3,442.0 | Dominated |
| food, afternoon, youth only | 230 | 1761 | 15.0 | \$18.1 | \$4,161.0 | Dominated |
| Wi-Fi, food, afternoon | 260 | 1791 | 17.0 | \$16.4 | \$4,251.4 | Dominated |
| Wi-Fi, food, afternoon, youth only | 352 | 1883 | 23.0 | \$19.2 | \$6,758.1 | \$43.7 |

**Table 3:** Clinic B: Cost-effectiveness of interventions to increase youth uptake of HIV and contraceptive services

|  | Additional youths | Total youths | % | Cost per patient | Total Cost | ICER |
| --- | --- | --- | --- | --- | --- | --- |
| Wi-Fi | 49 | 1857 | 2.7 | \$10.6 | \$522.7 | Weakly dominated |
| youth only | 5 | 1813 | 0.3 | \$116.5 | \$611.1 | Dominated |
| Wi-Fi, youth only | 163 | 1971 | 9.0 | \$7.0 | \$1,141.9 | \$7.0 |
| afternoon | 15 | 1823 | 0.8 | \$77.6 | \$1,150.6 | Dominated |
| Food | 41 | 1849 | 2.3 | \$30.8 | \$1,254.8 | Dominated |
| food, youth only, | 145 | 1953 | 8.0 | \$11.4 | \$1,653.6 | Dominated |
| afternoon, youth only | 125 | 1933 | 6.9 | \$13.8 | \$1,721.0 | Dominated |
| Wi-Fi, food | 199 | 2007 | 10.7 | \$9.0 | \$1,785.6 | Weakly dominated |
| Wi-Fi and afternoon | 168 | 1976 | 9.3 | \$7.5 | \$1,253.7 | Dominated |
| food, afternoon | 163 | 1971 | 9.0 | \$14.8 | \$2,413.4 | Dominated |
| Wi-Fi, food, youth only | 307 | 2115 | 17.0 | \$8.1 | \$2,490.5 | \$9.3 |
| Wi-Fi, afternoon, youth only | 271 | 2079 | 15.0 | \$8.5 | \$2,293.3 | Dominated |
| Wi-Fi, food, afternoon | 307 | 2115 | 17.0 | \$12.7 | \$3,914.4 | Dominated |
| food, afternoon, youth only | 271 | 2079 | 15.0 | \$11.2 | \$3,026.0 | Dominated |
| Wi-Fi, food, afternoon, youth only | 416 | 2224 | 23.0 | \$9.8 | \$4,059.7 | \$14.5 |

The addition of subsidized food had the third highest cost per additional adolescent utilising HIV and contraceptive services at US$31.5 for clinic A and US$30.8 for clinic B. This was because we cost the food to benefit all adolescents utilising services but the cost would be borne only by the additional youth utilising services. Provision of services in the afternoon outside school hours has the next highest cost per additional adolescent utilising services (US$78.5 per additional adolescent at clinic A and US$77.6 at clinic B). Providing youth only services would be the costliest per additional youth accessing HIV and contraceptive services with costs for clinic A at US$202.9 and clinic B at US$116.5. Combining all modifiable attributes was US$19.20 for clinic A and US$9.76 for clinic B per additional adolescent utilising services.

Figure 4 shows all non-dominated interventions scenarios for the two clinics, ranked in order of decreasing cost effectiveness. The most value for money is gained by implementing the interventions from the left to the right until the budget is exhausted (28). For both clinics, the ICER for youth only services and Wi-Fi was the best. The ICER’s for clinic A range from US$9.78 for (Wi-Fi and youth only services) and US$43.7 for a combination of all four modifiable attributes. The same order is retained with regards to ICER’s for clinic B, though with lower incremental costs-US$7.01 for (WI-FI and youth only services) and US$14.5 for all four modifiable attributes combined.

**Figure 4:**
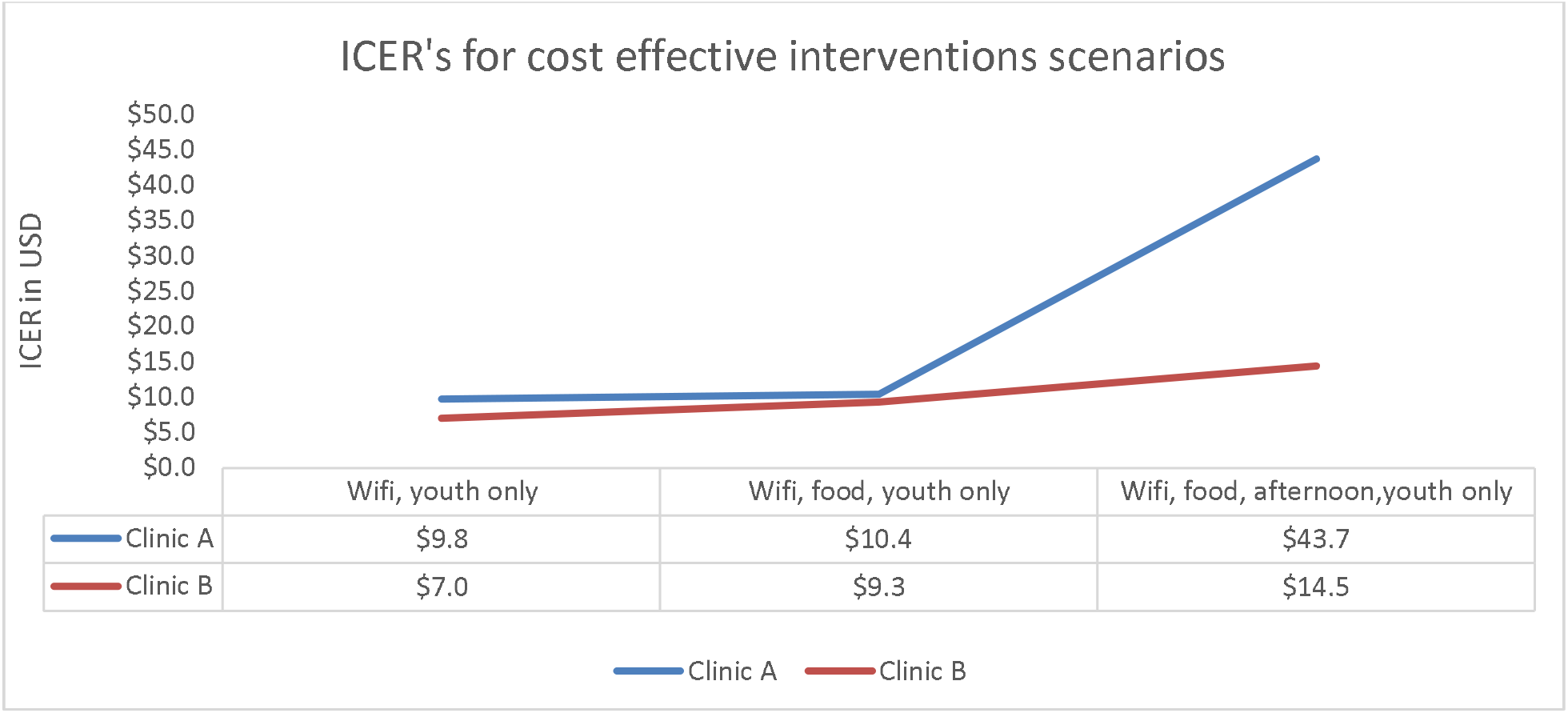
Incremental cost-effectiveness ratios on the cost-effectiveness frontier

## Discussion

We have described a methodology to estimate cost-effectiveness of potential service delivery mechanisms-for HIV and contraceptive services for youth-utilising results of a discrete choice experiment. Typically, cost-effectiveness analyses only come at the end of costly and time-consuming trials or large-scale observational studies. This type of pre-intervention assessment of potential cost-effectiveness of a set of interventions based on the stated preferences of the target audience and an estimate of costs of each intervention can assist in guiding more efficient trial planning-such that only the arms that are expected to be on the cost-effectiveness frontier are included in a trial design. Results could also be used to directly guide programme planning and scale-up where appropriate. The strength and innovation of this study lies in the use of DCE data to calculate potential changes in uptake of services from the current standard of care if different interventions were implemented, and also the use of cost data from clinics to calculate incremental costs of adding interventions. The use of data from DCEs to make projections on uptake of services to inform cost effectiveness analysis is valuable for intervention trials where resources do not exist to do the trial and obtain actual uptake data (revealed preferences) (30–32).

The results from the first analysis applying this novel methodology were able to determine the set of non-dominated strategies for increasing uptake of services, and determine the order with which they should be implemented across the cost-effectiveness frontier.

Friendliness, privacy and confidentiality are among the strongest drivers of choice for use of HIV and contraceptive services among adolescents who participated in the DCE used in this approach. We stratified the preferences identified from the DCE into modifiable and non-modifiable preferences. Friendliness and confidentiality were considered non-modifiable preferences-meaning that interventions to address these preferences are currently not well-defined (and therefore not cost-able). The DCE results and uptake projections show, however, that the non-modifiable factors are the most impactful and would provide the biggest increase in uptake-if an intervention to address these factors could be well-defined and costed. Addressing staff attitudes with regards to friendliness and confidentiality is likely a cost-effective strategy(33,34) as the impact is likely to go beyond just utilization of HIV and contraceptive services for adolescents into other health and educational outcomes (35,36).

The effectiveness of value-added services like access to free or affordable food, Wi-Fi and cash transfers has been evaluated before (7,34). Although cost-effective, it is important to be aware of their full impact and where this impact may plateau and new strategies must come in. Wi-Fi and youth-only services cost an incremental $7.01 - $10.62 per additional adolescents expected to access the healthcare facility but on their own are only expected to increase adolescent utilization of health services by 9%. Should resources allow, additional considerations would be, first: Wi-Fi and youth-only services, second: the addition of subsidized food and third: addition of afternoon clinic hours. Our results show that the order that we must implement interventions is the same between the two clinics and that altering the size of the clinic and other parameters may change the cost per additional adolescent utilising services.

This study was subject to a number of limitations. Firstly, we were unable to cost the non-modifiable preferences-although these produced the largest impact regarding uptake of services. Ideally, more work needs to be conducted to investigate ways to increase healthcare provider attitude. Behavioural economics informed interventions or nudges provide a potential way to influence provider attitude and behaviour (37). Studies that investigate possible costs and effectiveness of interventions that are geared towards improving provider attitudes towards adolescents and their access to HIV and contraceptive services would be required to effectively evaluate the cost-effectiveness of such interventions in this modeling framework. Second, projections from the DCE are theoretical as they are based on stated preferences as users are not observed (38). Results from DCEs have, however, been used in other studies to project uptake of services or products (31,39). A DCE forms a reliable evidence base for such projections. Third, our study does not look at the heterogeneity of preferences and therefore differentiated impact depending on the underlying population. Relative expected cost-effectiveness might differ based on the composition of the underlying population. A DCE conducted across multiple different underlying populations or in different geographies would address this limitation of the present application of this novel methodology.

## Conclusions

Patient-centred care has been expounded as the ultimate goal of health care service provision but is rarely a reality. The work described here might provide an approach to meet that need whilst considering the resource limitations that often exist particularly in low-resource settings. Policy decision making requires consideration of both patient preferences and cost-effectiveness information our study utilizes the DCE methodology to infer both. In instances where funding or time do not allow, the proposed methodology provides one mechanism to understand the possible cost-effectiveness of different implementation scenarios. The results from this type of analysis can be useful in either guiding implementation-if no plan exists for further study- or could be used to guide study design for empirical cost-effectiveness studies. Follow up work could look at acceptability, feasibility, and effectiveness of interventions built from these scenarios on uptake of HIV services by young people in different settings.

## Data Availability

All data produced in the present study are available upon reasonable request to the authors

## Authors’ contributions

[Mandatory]

CG and BEN conceptualized and designed the study. CGM conducted the analysis. CG prepared the original draft. CG, BEN, SP, LL, CAR and AM reviewed and edited the draft.

## Acknowledgements

[Mandatory]

## Funding

This study was made possible by the generous support of the American People and the President’s Emergency Plan for AIDS Relief (PEPFAR) through US Agency for International Development (USAID) under the terms of Cooperative Agreements AID-674-A-12-00029 and 72067419CA00004 to Health Economics and Epidemiology Research Office. LL was supported by the National Institute of Mental Health of the National Institutes of Health under grant number K01MH119923. The contents are the responsibility of the authors and do not necessarily reflect the views of the NIH, PEPFAR, USAID or the United States Government. The funders had no role in the study design, collection, analysis and interpretation of the data, in manuscript preparation or the decision to publish.

Disclaimer:

### Additional files

[Optional]

Additional file 1: Title of additional file

Information on file format. Brief description of file content.

## List of abbreviations

[Optional]

## Sites

- South African non-profit organization providing Comprehensive PHC services provided by nurses and doctors. DOH support-Accesses public-sector drugs and laboratory tests.
- Government, primary health clinic (PHC)

Sites reported similar mean age, days in care, and number of visits.

## Costing

- Resource use was calculated from the perspective of the provider, the public health system.
- All costs were updated to 2019 public-sector prices and salaries

### Summary of costs, assumptions and sources of cost data for each scenario

| Cost categories | Methods | Assumptions | Source |
| --- | --- | --- | --- |
| Total PHC headcounts were calculated using monthly headcounts over a period of 12 months (2018) |  |  |  |
| **Staff (clinical and non-clinical)** | top-down approach for staff | Additional youths would bear the cost of all additional staff at the facility as a result of intervention | Data collected for Long et al(1) |
| **Consumables/supplies** | Top-down. Calculated using monthly expenditure on supplies. This was shared among all PHC patients. (Total spent on consumables/total number of patients). | Maintained the per patient cost of consumables at the clinic level for the new youth patients obtained. | Data collected for Long et al(1) |
| **Overheads** | Overhead costs were calculated through a top-down approach. Additional overhead costs, i.e. extra space for youth only were all shared by additional youth. | Separate space the size of a single counselling room at the facility was allocated for youth only services.<br>Overheads were calculated by multiplying total overhead expenditure by proportion of new youths over total PHC count. | Data collected for Long et al(1) |
| **Assets** | Bottom up inventory of all assets in clinic and shared among all patients. | Shared the total cost of assets amongst all patients as no new assets would be necessary for intervention. | Data collected for Long et al(1) |
| **WIFI** |  | **WIFI** - Project Isizwe (Free WIFI for Tshwane clinics average costs). Costs of supply to a clinic for 1 month shared by all youths. | Data collected for Long et al(1) and (2) |
| **Subsidized food** |  | All youth presenting received subsidised food but additional youths bore the cost. A total of ZAR 10 was cost for each adolescent patient. | **Costs from (3)** |

